# Examining Gaps in Institutional Policies for Clinical Genomic Data Sharing: A Cross-Jurisdictional Study

**DOI:** 10.64898/2026.03.09.26347975

**Authors:** Zhaoping Ju, Yunhe Xue, Abby Rud, Juliann M. Savatt, Jordan Lerner-Ellis, Heidi L. Rehm, Yann Joly, Diya Uberoi

## Abstract

**Background:** The sharing of data generated through the course of clinical genetic and genomic testing without explicit patient consent is increasingly important for timely diagnosis and treatment. While many jurisdictions permit the sharing of identifiable data for direct patient care, institutional policies vary in how clearly they specify key elements. When do policies permit sharing of data without explicit consent? What data types may be shared, with whom, and under what safeguards? Greater clarity around these elements may support responsible data sharing while balancing timely care with transparency and appropriate protections.

**Methods:** We conducted a qualitative content analysis of data-sharing and privacy policies from 33 clinical genomic institutions across 17 jurisdictions. Using a predefined analytical framework, we assessed how policies document key governance elements relevant to sharing without explicit consent. Two independent reviewers extracted information about clinical contexts, data types, justifications, and protections, documenting areas of inconsistency across institutions.

**Results:** Although 70% of institutions described circumstances permitting data sharing without explicit consent, most policies did not clearly define the scope or governance of such sharing. Policies also rarely distinguished clinical from research or secondary use and inconsistently specified privacy and security safeguards. While sharing was commonly justified for clinical care (78.3%) or testing services (43.5%), recipient roles, access conditions, and onward-sharing expectations were often left undefined.

**Conclusion:** This uneven documentation could make it difficult for clinical teams, laboratories, and institutional decision-makers to identify and justify key decisions about what is permitted and under what conditions. A guidance framework specifying core policy elements and corresponding protections could help institutions communicate their governance choices more clearly while supporting more comparable baseline practices for responsible data sharing across settings.

## Introduction

As genomic medicine becomes embedded in routine clinical practice, sharing data generated as part of clinical genetic and genomic testing without explicit patient consent has become increasingly important for timely care^1^. By enabling aggregation of case-level evidence across institutions and supporting reinterpretation as new information emerges, cross-institutional sharing can strengthen clinical interpretation, improving variant classification and refining gene–disease and genotype-phenotype relationships relevant to diagnosis and treatment. This exchange is often facilitated through interpretation and matching platforms such as ClinVar and the Matchmaker Exchange (MME). ClinVar submissions, for example, have been used to uncover differences in variant classification and to help laboratories identify where a variant has been observed, enabling follow-up requests for case-level observations that support more accurate diagnosis^2,3^. MME similarly supports rare disease diagnosis through a federated platform that enables laboratories and clinicians to share and match candidate genes and variants across institutions, allowing the aggregation of evidence to inform new gene–disease discoveries and improve phenotypic characterization^4,5^.

However, while these resources have made cross-institutional exchange clinically valuable, they also make it difficult to rely on explicit patient consent for each instance of variant interpretation-related sharing. In practice, explicit consent is not typically obtained for exchanges through platforms such as ClinVar and MME, particularly when case-level information is shared without direct identifiers^6,7^. Because these exchanges recur over time and extend across institutions, mandating consent for each disclosure would be impractical and may delay interpretation by limiting access to comparative data and slowing collaboration^8,9^. This is particularly true when sharing requests are directed to clinical testing laboratories, which typically lack direct patient relationships and patients’ up-to-date contact information^2,3^.

Given these realities, many jurisdictions, including the United States^10^, the European Union^11^, the United Kingdom^12,13^, Australia^14^, and Canada^15–17^ (through provincial health privacy statutes), allow the sharing of identifiable patient data for direct patient care without requiring explicit consent for each disclosure. When patient data are exchanged within strictly governed clinical settings—utilizing, for example, encryption, pseudonymization, and secure access-controlled environments—the likelihood of re-identification and misuse can be reduced^18–20^. However, when institutions attempt to operationalize these permissions, significant variation can emerge in how laws and policies are interpreted and applied. For example, some jurisdictions permit disclosure of identifiable clinical information to clinicians or laboratories outside a patient’s immediate care team for expert consultation or variant interpretation^12,13^. In practice, however, these permissions are sometimes applied more restrictively, limiting disclosure to pseudonymized data even when more detailed case-level phenotypic and clinical information could support variant classification and timely diagnosis^21,22^.

These inconsistencies point to a fundamental governance challenge. In implementing jurisdictional permissions, institutional policies may not clearly specify how sharing decisions should be made or what limits and safeguards apply, reducing transparency and making accountability harder to assess. This can leave clinicians, laboratories, patients, and other stakeholders uncertain about the practical boundaries of permitted sharing and the rationale for particular decisions. Data sharing requires balancing timely access with appropriate privacy and security protections^23,24^. In clinical genomics, where interpretation and matching platforms make cross-institutional exchange routine and ongoing, this balance can be difficult to achieve. As data persist and circulate through public and controlled-access repositories (e.g., ClinVar, Matchmaker Exchange, Trusted Research Environments), intended uses may evolve and the boundary between clinical and research use can blur. At the same time, data types vary in identifiability and re-identification risk, and safeguards vary in scope, underscoring the importance of examining how institutional policies specify the conditions in which sharing proceeds in the absence of explicit consent^23,25^.

Standardized guidance could enable institutions to make transparent and comparable decisions about when and how to share clinical genomic data while maintaining trust and protecting patient autonomy. This is particularly important as organizations like the Global Alliance for Genomics and Health (GA4GH), an international standard-setting institution that brings together genomic scientists, clinicians and policy-makers from around the world, work to develop frameworks that support responsible data governance^26^. Drawing on input from contributors engaged in GA4GH clinical data sharing discussions and the Clinical Genomics Laboratory Community (CGLC), we examine the ethical and legal considerations institutions and laboratories must address to clarify clinical data sharing policies and practices^27^4.

This study provides an empirical analysis of institutional policies to identify where greater clarity is needed. Examining policies from 33 clinical genomic institutions across 17 jurisdictions, we characterize institutional approaches to sharing data generated through clinical genetic and genomic testing without explicit consent and identify where policies lack specificity regarding scope, purpose, safeguards, and transparency. We then describe how institutions document these elements when sharing such data, drawing on a qualitative review of institutional documents and genomics data governance literature. Ultimately, we envision that these findings can help inform cross-institutional dialogue about baseline policy elements that enable transparent, accountable clinical data sharing practices.

## Methods

### Selection Criteria

We conducted a qualitative content analysis of institutional documents describing clinical genomic data sharing and/or patient privacy practices across multiple jurisdictions. Institutions were purposively selected to capture regional diversity and variation in clinical genomics service models and governance contexts. Because inclusion required publicly accessible policy materials, geographic representation depended on the availability of institutional documentation.

Potential institutions were identified through targeted web searches of publicly available resources describing clinical genomics providers, including health-system listings. A small number were also identified through suggestions by GA4GH members. For each institution, we systematically searched institutional websites to obtain documents describing data sharing, privacy, and/or permitted disclosures of patient information. When relevant materials were not available online, members of the research team requested them through correspondence with institutional representatives.

Institutions were eligible for inclusion if they offered clinical genomics care or genetic or genomic testing services and had at least one publicly accessible data sharing policy available through their website or through direct correspondence with institutional representatives. All relevant documents from a given institution were treated as an institutional policy set for coding. Where policies were not available in English, relevant sections were translated for coding using online translation tools. When policies did not enumerate data types in scope, we used publicly available institutional testing offerings and service descriptions to characterize the data likely covered by the policies. Specifically, we reviewed institutional service pages to identify the types of testing offered and clinically relevant information used in interpretation, such as phenotype-driven services and family-based testing.

### Analytical Framework & Coding Approach

To identify gaps in institutional guidance on clinical genomic data sharing without explicit consent, we qualitatively analyzed policy content across six key dimensions. These included: (1) data sharing consent requirements and criteria for waiving consent, (2) data types and scope (including any sensitivity distinctions), (3) stated justifications/purposes for sharing (including how policies distinguish clinical from secondary or research use), (4) intended recipients, (5) documented safeguards and accountability mechanisms, and (6) patient-facing transparency and consent mechanisms (including opt-out). These criteria were based on a narrative review of scholarly literature on the ethical and governance aspects of clinical genomic data sharing and were refined through an initial review of three institutional policies.

Two researchers (Z.J. and Y.X.) independently reviewed and coded each policy document according to the dimensions above. Discrepancies in coding were resolved through discussion and consensus. When needed, a third reviewer (D.U.) finalized coding decisions. Data were organized and analyzed in Microsoft Excel to facilitate cross-institutional comparisons. We used descriptive statistics, including counts and percentages of institutions with specific policy features, and graphical representations to identify commonalities and differences across institutions.

## Results

### Demographics

Of the 33 institutions included in our analysis, 10 (30.3%) were located in Europe, eight (24.2%) in Asia, eight (24.2%) in North America, six (18.2%) in Oceania, and one (3.0%) in South America **(Figure 1)**.

**Figure 1:**
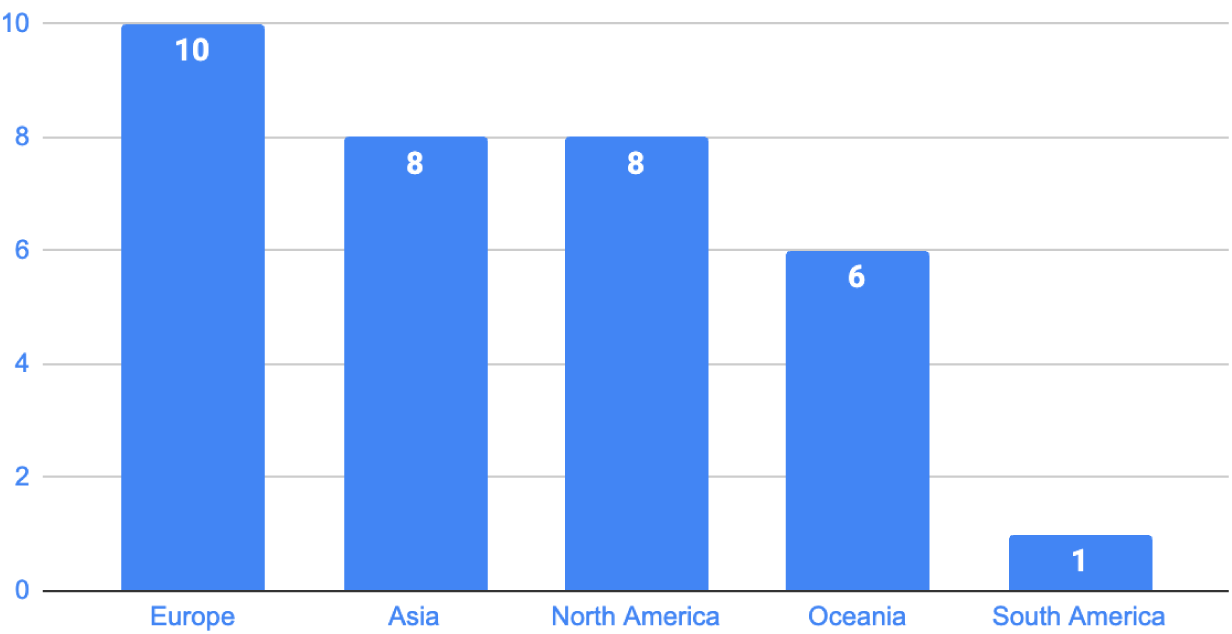
Geographic distribution of selected clinical genomic institutions

The dataset included a nearly even split between publicly funded (n = 16, 48.5%) and privately operated (n = 17, 51.5%) organizations. Most institutions provided both direct clinical care and in-house genomic testing (n=19, 57.6%), while the remainder either offered clinical care without genomic testing (n=3, 9.1%) or functioned as testing laboratories without direct patient care services (n=11, 33.3%).

We found that policies varied widely in how clearly they described the circumstances under which clinical genomic data may be shared without explicit consent. Below, we report findings across the six policy dimensions defined in the Analytical Framework and Coding Approach. For each dimension, we report descriptive counts and observed patterns to support cross-institutional comparison.

### Consent requirements and criteria for waiving consent

Among the 33 institutional policies reviewed, 23 (23/33, 69.7%) described circumstances permitting some sharing without explicit consent, nine (9/33, 27.3%) required explicit consent in all cases, and one (1/33, 3.0%) prohibited third-party sharing under any circumstance (**Figure 2**).

**Figure 2:**
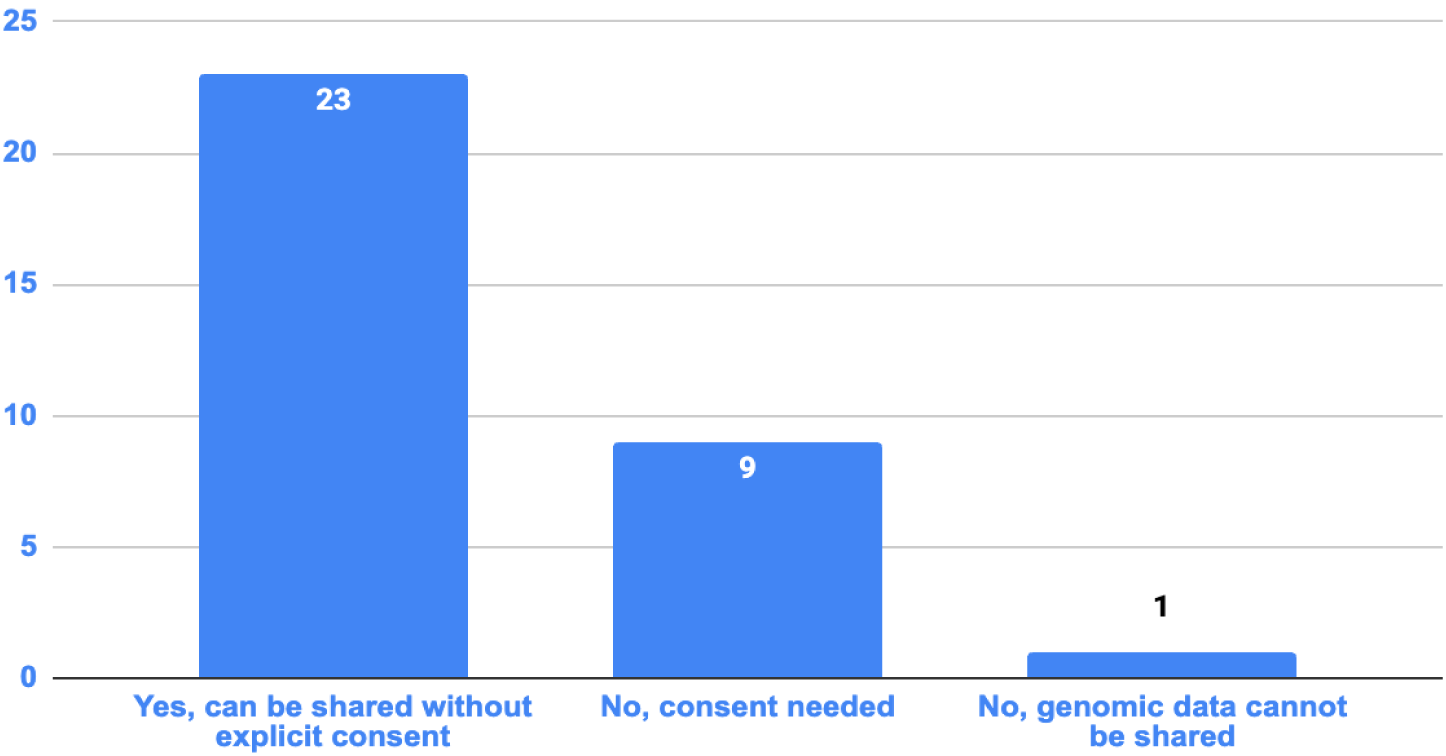
Consent practices for clinical genomic data sharing

Even among the 23 institutions that permitted sharing without explicit consent in certain circumstances, policies seldom described how decisions to disclose without explicit consent should be made or documented. Most did not specify decision criteria (e.g., when a waiver is appropriate), thresholds (e.g., what conditions must be met), or procedural requirements (e.g., who authorizes disclosure, what documentation is required, or whether any review process applies). Instead, policies often relied on permissive statements (e.g., allowing disclosure “without consent”) without explaining how such decisions should be justified or recorded. This gap is illustrated by one policy, which stated that “Information will not be disclosed…unless this request is authorised by a court of law, or you have given specific permission,” but did not indicate who makes that determination, what supporting documentation is required, or whether any review applies (Inst. #30).

### Data types and scope (including sensitivity distinctions)

Policies also did not specify what types of genomic or clinically relevant data were covered, or whether any sensitivity distinctions were defined. Given this lack of clarity, we characterized likely policy scope using institutional test offerings and service descriptions.

Across the 23 institutions that permitted sharing without explicit consent, all offered both somatic and germline testing (23/23, 100%), including tumour profiling and/or inherited disease testing. Most offered targeted gene panels or genome-wide sequencing of genomic data (15/23, 65.2%). Mitochondrial DNA testing was offered by ten institutions (10/23, 43.5%). RNA-based diagnostics were offered by 11 (11/23, 47.8%), and epigenetic tests (including DNA methylation profiling) by nine (9/23, 39.1%) **(Figure 3A)**.

**Figure 3:**
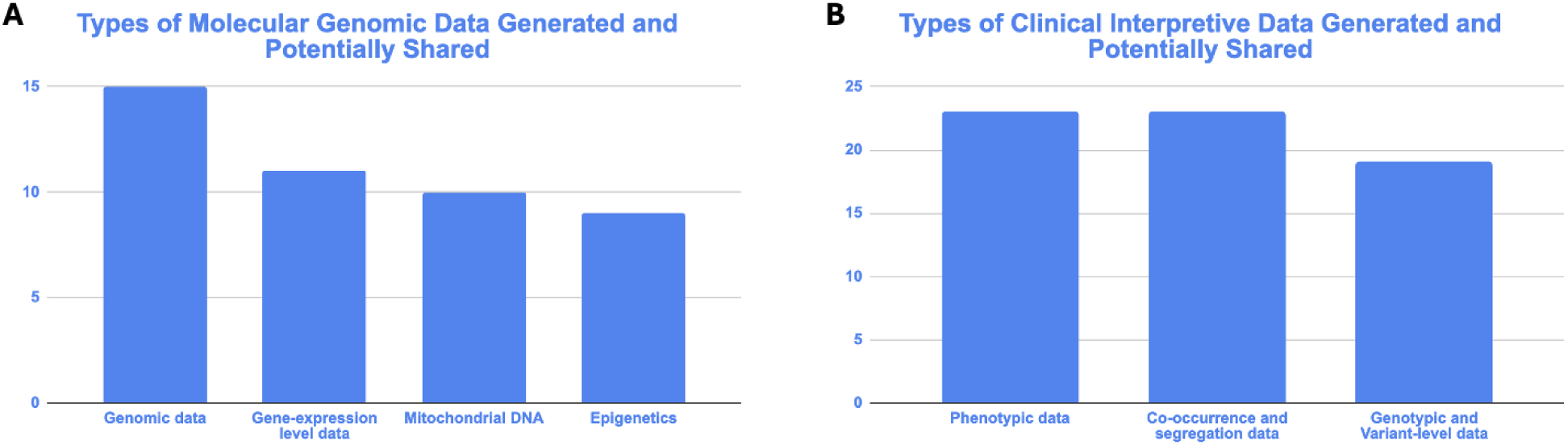
Types of data generated and potentially shared

Beyond molecular data, institutions’ test offerings and service descriptions indicate that clinical genomic services routinely rely on phenotypic and familial information used alongside genomic findings. All 23 institutions referenced clinical services requiring phenotypic information (23/23, 100%), such as phenotype-driven interpretation or clinician-provided diagnostic details, and all offered family-based testing (23/23, 100%), including trio testing or segregation analysis, which can generate co-occurrence and segregation information. While phenotype-driven services and family-based testing were referenced across all 23 institutions (23/23, 100%), variant classification activities or reports containing variant-level information were explicitly referenced by 19 (19/23, 82.6%) (**Figure 3B**).

Taken together, these service offerings suggest that the practical scope of “data” potentially covered by institutional policies is potentially broad. Yet, despite this breadth, policies generally did not distinguish among these categories when describing sharing permissions, nor did they specify how different data types should be handled based on relative data sensitivity or re-identification risk.

### Rationale and purposes for sharing

Policies that permitted sharing without explicit consent typically explained why sharing should occur, but were often less specific about how those stated purposes translate into operational boundaries, including what conditions apply, the data in scope, and whether different purposes are subject to different requirements. Most commonly, policies framed sharing as supporting the provision of clinical genomic care, including information exchange among healthcare professionals to support diagnosis or treatment (18/23, 78.3%). A related justification was the provision of clinical genomic testing services, including the exchange of data and/or samples with laboratories (10/23, 43.5%). While most policies emphasized clinical care or testing services, some also invoked broader legal bases, including compelling legitimate interests or legal obligations (10/23, 43.5%) and vital interests, such as emergency or life-threatening situations (6/23, 26.1%) (**Figure 4**). For example, one policy indicated that disclosure may be permitted “where there is a strong public interest in disclosure that outweighs the public interest in maintaining confidentiality,” but did not specify how that balancing determination should be made (Inst. #1).

**Figure 4:**
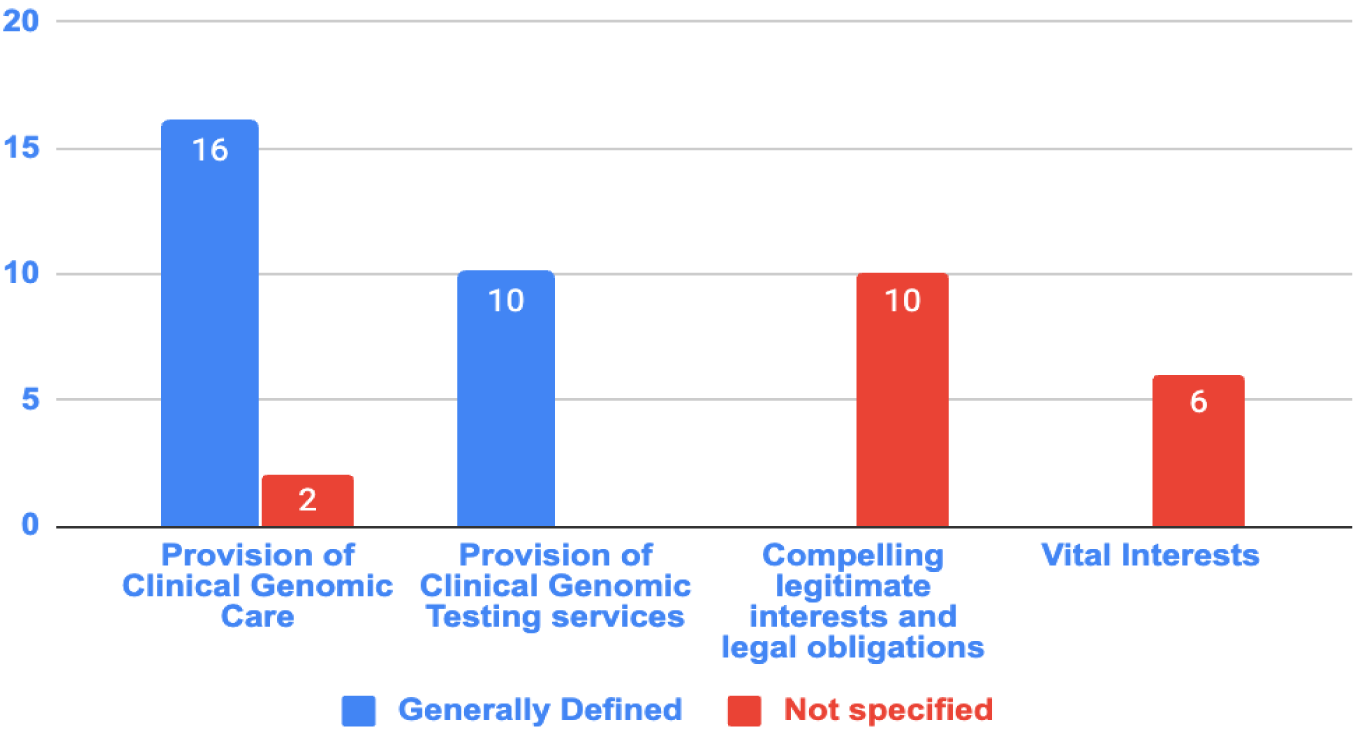
Purposes and recipient categories for sharing without explicit consent in institutional policies. *. Note-categories are not mutually exclusive. Percentages were calculated using the number of policies referencing each purpose as the denominator.

Because stated purposes are often expressed through recipient categories and access conditions, we examined how policies described who may receive data and under what access arrangements.

### Intended recipients and access contexts

Institutional policies permitting the sharing of clinical genomic data without explicit consent consistently described authorized recipients using broad or nonspecific terms. Recipient descriptions typically referred to general categories such as healthcare professionals, laboratories, clinical partners, or authorized third parties, without further role-based or credential-specific clarification. (**Figure 4).**

When policies referenced sharing for the provision of clinical genomic care, recipient descriptions were either not specified (n = 2/18) or generally referred to data exchange among healthcare providers involved in diagnosis or treatment (n = 16/18), such as referring physicians or genetic counselors. Policies invoking the provision of clinical genomic testing services typically referred to laboratories responsible for conducting or interpreting genetic tests (n = 10/10) (**Figure 4**). Notably, none of the policies explicitly enumerated recipient categories or described authorization mechanisms governing access to shared data.

Similarly, when sharing was justified by compelling legitimate interests and legal obligations (n = 10/10) or vital interests (n = 6/6), recipient descriptions were typically not clarified (**Figure 4**). Downstream access conditions and onward-sharing limitations were also rarely specified. Although such sharing may be assumed to occur within a “circle of care,” most policies did not specify when sharing extends beyond that circle or how recipient boundaries are determined.

### Secondary uses beyond direct care

Even when policies referenced secondary uses beyond direct clinical care, they often did not make clear whether (or how) those uses were governed differently from direct care. Nineteen institutions referenced data retention or use beyond immediate clinical encounters (19/23, 82.6%), and 21 referenced research or secondary uses of patient data in some capacity (21/23, 91.3%). For example, one policy states that “We may use your health information for administrative,..financial and…quality improvement activities,” but did not specify whether these secondary uses were subject to different conditions than sharing for direct clinical care (e.g., different consent requirements, governance steps, or data handling expectations) (Inst.#19). While secondary or research use was frequently mentioned, only seven of these 21 institutions (33.3%) clearly distinguished between sharing for direct patient care and sharing for secondary or research purposes, for example by specifying different consent requirements, data identifiability expectations, or governance mechanisms (**Figure 5**).

**Figure 5:**
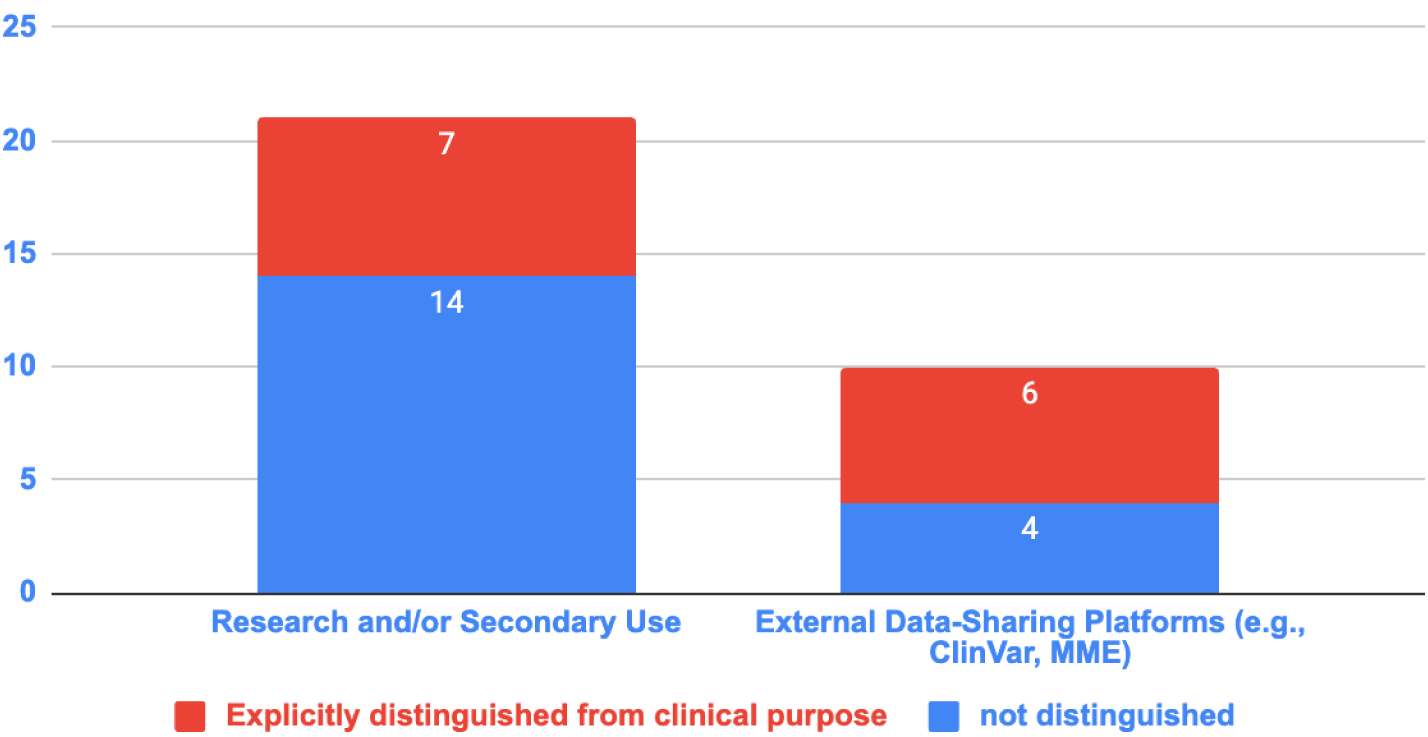
How policies describe sharing and use beyond direct clinical care

This ambiguity is especially salient when policies referenced sharing through publicly accessible databases and interpretation or matching platforms used in clinical genomics, where information may be retained and accessed beyond the primary clinical interpretation purpose. Ten institutions (10/23, 43.5%) named databases or platforms such as ClinVar^7^ or the Matchmaker Exchange^5^. However, among these ten, only six (60.0%) provided platform-specific guidance indicating how submissions are made, whether they are handled differently from other disclosures described in policies, or what happens after submission, including follow-up inquiries **(Figure 5).**

### Documented safeguards and accountability mechanisms

Given the frequency of secondary uses and the limited clarity on how they were governed, it became important to examine what safeguards and accountability mechanisms policies specified. Across the 23 institutions that permitted clinical genomic data sharing without explicit consent, most policies did not specify any concrete safeguards associated with such sharing. Sixteen institutions did not reference any specific technical, administrative, or procedural security or privacy measures in their formal policy documents (16/23, 69.6%).

Among the seven institutions that did describe safeguards, all referenced at least one technical security measure (7/7), and four also described administrative or organizational measures (4/7), such as staff confidentiality training and/or internal oversight procedures (**Figure 6A**). When technical measures were mentioned, they typically addressed secure storage and transfer, most often through encryption and access controls. Three institutions also explicitly referenced de-identification or pseudonymization as part of their technical safeguards (3/7, 42.9%).

**Figure 6.**
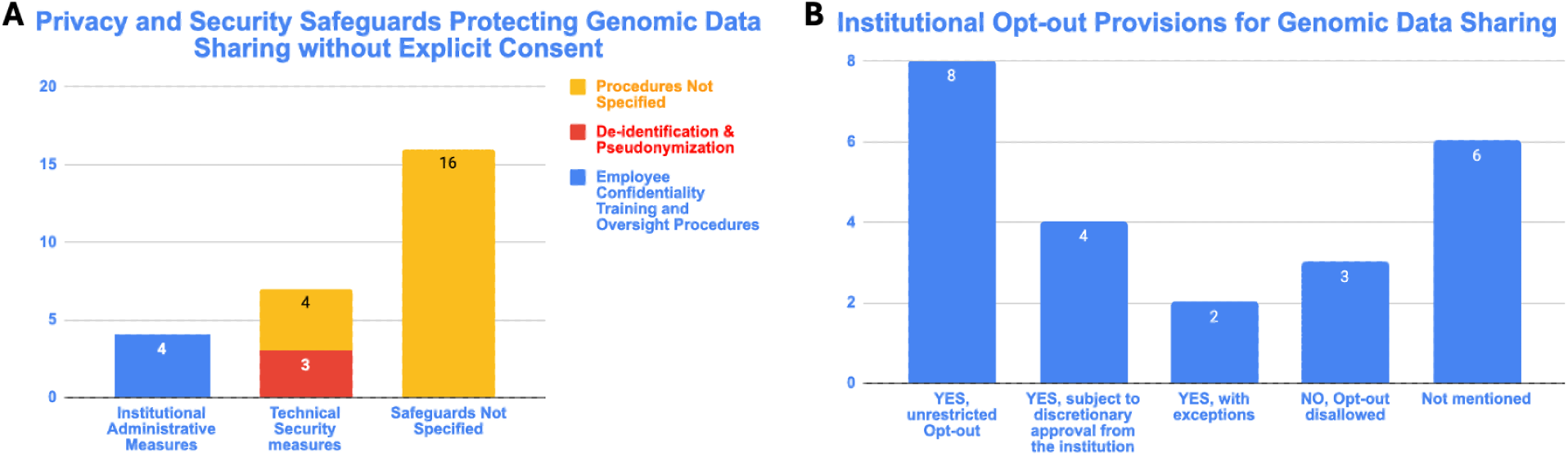
Safeguards and patient control mechanisms described in institutional policies

Even where safeguards were mentioned, the level of specificity was highly variable. Some policies listed general categories of protections, but did not clarify scope, including which users and what the protections cover, and what accountability steps accompany sharing without explicit consent. As a result, there was no consistent approach across institutions to documenting the minimum safeguards associated with clinical data sharing.

### Opt-Out & Consent based mechanisms

Beyond privacy safeguards, some institutions described consent-based mechanisms intended to preserve autonomy when sharing occurs without explicit consent. In particular, several policies presumed agreement to data sharing unless patients affirmatively declined through an opt-out process.

Among the 23 institutions that permitted sharing without explicit consent, 14 (14/23, 60.9%) described some mechanism for patients to opt-out, though the accessibility and strength of these mechanisms varied. Of these 14 institutions, eight (8/14, 57.1%) allowed patients to opt out without restrictions. By contrast, four (4/14, 28.6%) required institutional approval before an opt-out request would be honored, and two (2/14, 14.3%) permitted opt-out but provided legal or contractual exceptions that could limit its implementation **(Figure 6B)**.

Conversely, only three institutions (3/23, 13.0%) explicitly prohibited any opt-out option, and six (6/23, 26.1%) did not address opt-out at all **(Figure 6B)**.

Because transparency and control mechanisms can shape who benefits and who may bear the burdens of clinical data sharing, policies’ silence on equity is consequential. In this context, we examined whether policies explicitly addressed equity considerations associated with consent waivers and opt-out mechanisms. Notably, none of the 33 institutional policies reviewed included explicit language addressing equity, health disparities, or the differential impact of clinical genomic data sharing on underrepresented populations. Policies also did not reference mechanisms to assess or mitigate potential inequities arising from consent waivers, or unrestricted data-sharing practices, nor did they include provisions aimed at ensuring equitable benefit from genomic data use.

## Discussion

Our analysis of data-sharing and privacy policies from clinical genomic institutions suggests that while legal frameworks may permit clinical genetic and genomic test-related data sharing without explicit consent, institutional documentation varies in how clearly it defines key operational boundaries. Across the institutions examined, most policies described or permitted sharing without explicit consent for direct patient care, but few specified the conditions or procedural requirements needed to govern such sharing (e.g. decision criteria, authorization, or review expectations). Because this study analyzes the content of policies rather than their implementation, our findings should be interpreted as documenting gaps and inconsistencies in institutional guidance rather than how those policies are carried out in practice. Together, these patterns highlight opportunities to articulate clearer baseline policy elements that support consistent, transparent decision-making about sharing clinical genomic data.

Many policies did not clarify what types of genomic or clinically relevant information were covered or whether safeguards should differ by the sensitivity of the data shared. While some policies named broad recipient categories (e.g., healthcare providers, laboratories, authorized third parties), they rarely specified access conditions, onward-sharing limits, or documentation expectations, making the practical boundaries of permitted sharing difficult to interpret. In practice, this ambiguity can lead to inconsistent disclosure decisions across teams and make it difficult to document the rationale for sharing^28,29^. In rare disease contexts, it can also slow the cross-institutional sharing needed to classify variants and reach diagnoses^30,31^.

Even when policies permitted sharing, safeguards and consent-based mechanisms were unevenly documented. Only seven of the 23 institutions permitting clinical data sharing referenced any form of technical or administrative safeguards, such as encryption, access controls, or staff confidentiality training. Patient control mechanisms were also inconsistent. Some institutions described opt-out options, but their scope varied, with some institutions permitting unrestricted opt-out and others limiting or omitting it altogether. While opt-out options can give patients more autonomy over how their data are shared, managing these options requires laboratories to track patient preferences and apply them consistently across disclosures, which can be operationally burdensome. Moreover, given the complex, context-dependent trade-offs involved in data sharing, patients may not have sufficient information or support to choose options that are consistent with their desires for balancing potential benefits and harms^32–34^. Standardized best practice guidance and educational resources could therefore help institutions implement opt-out options and related controls in ways that protect patient autonomy while supporting timely, high-quality care^33,35^.

Policies also did not indicate whether protections should vary with the sensitivity of the data shared. Variant-level data accompanied by high-level phenotype information often carries lower privacy risks than comprehensive individual-level genome-wide sequencing data paired with detailed phenotype, as the former is typically less readily identifiable and, therefore, harder to link back to an individual^8,36^. At the same time, risk of re-identification can increase as additional case-level clinical or phenotypic detail is shared^8,36^. Research indicates that applying uniform safeguards across data types can create unnecessary barriers to sharing lower-risk data while failing to provide adequate protection for more sensitive information^37^. Yet institutional policies rarely made this distinction explicit. A proportionality approach would allow institutions to align safeguards and patient control mechanisms with the data’s identifiability, intended uses, and downstream reuse risks. This becomes especially important when sharing occurs through clinical interpretation and matching platforms, where disclosures can persist over time and be made available under open or controlled-access arrangements beyond the immediate context of care.

For example, many institutions share, or reference sharing genomic data to ClinVar and the Matchmaker Exchange (MME)^2,5^. While data sharing via these platforms can improve variant classification as interpretations evolve over time, submissions may persist and be accessed or reused for purposes beyond the original clinical episode of care^8^. Yet institutional policies often do not clearly specify how access is governed and what information is submitted. Some policies reference ClinVar or the MME, but do not indicate whether submissions are limited to variant-level assertions (e.g., gene/variant and classification) or include additional case-level information (e.g., phenotype or segregation data) that can be important for interpreting clinical significance. Without greater clarity on access conditions and submission content, institutions and users may be less able to anticipate downstream uses, apply appropriate safeguards, and explain sharing clearly to patients. These gaps could also have interpretive and privacy implications, particularly as expectations for open versus controlled access across jurisdictions may shift. The National Institutes of Health is currently consulting on a Draft Controlled-Access Data Policy and proposed revisions to its Genomic Data Sharing Policy^38^. If adopted, these changes could tighten expectations around submission scope and when genomic data may be shared openly versus through controlled-access mechanisms, with important implications for clinical interpretation platforms and repositories^38^.

More broadly, because data shared for clinical interpretation may also support a range of medical and scientific work, clear purpose boundaries are essential. We found that seven of the 21 policies that addressed secondary use (7/21, 33.3%) explicitly distinguished between sharing for direct patient care and sharing for research or other secondary purposes. This may be because it is increasingly difficult to separate these uses as we enter the learning healthcare system era with many countries and institutions launching biobanks and other methods that use patient data to directly inform improvements in care.^39^ Yet without clear purpose boundaries, institutions may have limited policy basis to apply appropriate safeguards and help patients understand the range of future uses. As data aggregation becomes increasingly central to modern genomic medicine, institutions will need clearer guidance on defining clinical versus research uses to maintain accountable and comprehensible governance.

By identifying where policies are less specific, we aim to highlight opportunities for reference materials and standards that can support responsible data sharing while preserving institutional flexibility. Institutions could benefit from more comparable documentation of key policy elements, including scope, safeguards, transparency, and purpose boundaries, as well as the extent to which equity considerations are addressed. Greater transparency around these elements can support cross-institutional learning even when institutions adopt different governance choices. In calling for more transparency, we do not imply that institutions should make identical policy choices. Some variation may be context-justified, as institutional differences may reflect distinct legal requirements and interpretations related to privacy and the use of health data across jurisdictions. In addition, how institutions justify sharing without explicit consent may differ depending on the health system in which they operate. In public health systems with broad access to clinically indicated services, sharing without explicit consent may be more readily framed as supporting direct patient benefit, whereas in more privatized systems, where access to downstream benefits may be less universal, claims of direct patient benefit may be harder to sustain^40,41^. Our findings instead point to the value of a shared core set of governance elements that institutions could use to articulate their policies more consistently, enabling more transparent comparison and more informed dialogue about where alignment is warranted and where context-specific differences should remain

## Limitations

While our findings provide insights into institutional approaches to clinical data sharing, certain methodological limitations should be acknowledged. Our analysis examined policies as written rather than their implementation, which limited our ability to assess whether safeguards, additional information, or practices not described in policy documents were used by data providers, repositories, or users. In addition, institutions were included only when relevant policies were publicly accessible or could be obtained through outreach, which may have favored institutions with more accessible policy documentation. Our sample, while geographically diverse across Europe, Asia, North America, Oceania, and South America, did not include institutions from all regions, thereby limiting the study’s generalizability. The study did not evaluate policies from a single user perspective, e.g., a laboratory test requisition versus a general hospital policy and therefore may not have captured all policy variation relevant to genetic data sharing. Despite these limitations, our study provides one of the first broad content analyses of institutional policies governing the sharing of clinical genomic data across jurisdictions.

## Future Directions

As clinical genomic medicine has advanced, cross-institutional sharing of clinical genetic and genomic test-related data has become increasingly important for diagnosis and care. Secure, responsible sharing without explicit consent depends, in part, on whether institutional policies clearly specify what data may be shared, with whom and for what purposes, and under what accountability mechanisms. Our analysis suggests that many policies remain insufficiently specific around these elements, which could make it harder for laboratories and other stakeholders to interpret governance choices that support transparent oversight.

Going forward, standard-setting organizations such as the GA4GH could build on this evidence to develop practical guidance that articulates baseline policy elements and sets out governance expectations. Because governance of clinical genomic data-sharing is often communicated through multiple policy sources written for different users (e.g., test requisitions, institutional privacy policies, and internal procedures), a next step could be to clarify how baseline elements should be expressed across this broader “policy set,” recognizing that no single document is likely to capture all relevant detail. This approach provides a foundation for mapping expectations onto common sharing scenarios, including lab-to-lab sharing and platform-mediated submissions, while clarifying how sharing without explicit consent is operationalized across contexts.

It can also help specify (i) which policy elements should be visible to which audiences including patients and laboratories, (ii) where documentation is necessary (e.g., in a test requisition form vs. institutional policy vs. internal procedure), and (iii) what minimum information is needed to support transparent oversight and patient communication even when institutions make context-justified governance choices. Together, these steps could support more comparable institutional practice and enable more productive cross-institutional dialogue about where alignment is needed.

## Data Availability

All data produced in the present study are available upon reasonable request to the authors

## Author Contributions

Zhaoping Ju and Diya Uberoi had full access to all of the data in the study and take responsibility for the integrity of the data and the accuracy of the analyses.

**Concept & Design:** Ju & Uberoi

**Original Draft:** Ju & Uberoi

**Data Curation & Analysis**: Ju & Xue

**Investigation**: Ju

**Writing, Review & Editing:** Ju, Xue, Rud, Savatt, Joly, Rehm, Lerner-Ellis, Uberoi

## Acknowledgements

We thank Erin Porter for her help editing an earlier version of this manuscript.

---

**Appendix 1:** Selected clinical genomic institutions included in the study.

| Selected Institutions |  |  |
| --- | --- | --- |
| Continent | Jurisdiction | Institution* |
| Europe | United Kingdom | 1. All Wales Medical Genomics Service (AWMGS)<br>(Website & Policy 1, Policy 2, Policy 3) |
|  | Ireland | 2. Children's Health Ireland - Department of Clinical Genetics<br>(Website & Policy 1, Policy 2, Policy 3) |
|  | Ireland | 3. Eurofins Clinical Genetics<br>(Website & Policy) |
|  | Germany | 4. CeGaT<br>(Website & Policy) |
|  | Germany | 5. The Zentrum für Humangenetik Tübingen<br>(Website & Policy (in DE)) |
|  | Finland | 6. Abomics<br>(Website & Policy) |
|  | Finland | 7. Blueprint Genetics<br>(Website & Policy) |
|  | Spain | 8. SJD Barcelona Children's Hospital<br>(Website & Policy) |
|  | Denmark | 9. Rigshospitalet - Department of Clinical Genetics<br>(Website & Policy) |
|  | Sweden | 10. Karolinska University Hospital<br>(Website & Policy) |
| Asia | Japan | 11. The University of Tokyo Hospital<br>(Website & Policy) |
|  | Japan | 12. Tottori University Hospital<br>(Website & Policy (in JA)) |
|  | Japan | 13. GENESISPRO<br>(Website & Policy) |
|  | South Korea | 14. 3Billion<br>(Website & Policy) |
|  | South Korea | 15. Asan Medical Center<br>(Website & Policy) |
|  | Hong Kong | 16. Hong Kong Sanatorium & Hospital<br>(Website & Policy**) |
|  | Singapore | 17. SingHealth (institutions)<br>(Website & Policy) |
|  | Thailand | 18. Ramathibodi hospital<br>(Website (in Thai) & Policy) |
| **North America** | United States | 19. BJC HealthCare (Washington University Physicians)<br>(Website & Policy) |
|  | United States | 20. Color Health<br>(Website & Policy 1, Policy 2) |
|  | United States | 21. Fulgent Genetics<br>(Website & Policy 1, Policy 2, Policy 3, Policy 4) |
|  | United States | 22. ARUP Laboratories<br>(Website & Policy 1, Policy 2) |
|  | United States | 23. Ambry Genetics<br>(Website & Policy) |
|  | United States | 24. Myriad Genetics<br>(Website & Policy 1, Policy 2) |
|  | Canada | 25. Sinai Health; Department of Pathology and Laboratory Medicine (Sinai Health System)<br>(Website & Policy) |
|  | Canada | 26. SickKids Hospital; Genome Diagnostics Laboratory, Hospital for Sick Children<br>(Website & Policy) |
| **Oceania** | New Zealand | 27. Genetic Insight<br>(Website & Policy) |
|  | New Zealand | 28. Genetic Health Service NZ<br>(Website & Policy 1, Policy 2, Policy 3) |
|  | Australia | 29. Victorian Clinical Genetics Services<br>(Website & Policy 1, Policy 2) |
|  | Australia | 30. Brisbane Genetics<br>(Website & Policy) |
|  | Australia | 31. Genetic Services of Western Australia (GSWA)<br>(Website & Policy) |
|  | Australia | 32. Genomic Health<br>(Website & Policy) |
| **South America** | Brazil | 33. Clinical Pathology, Hospital de Clínicas, Universidade Federal do Paraná (UFPR)<br>(Website & Policy (in portuguese)) |

*\*Policy and website embedded in hyperlink and date of last access is August 14, 2025.*

## Appendix 2: Definitions of clinical interpretive genomic data types

1. **Variant-level data:** Information about single variants, including the variant name, its clinical significance, the associated condition, and supporting evidence such as literature summaries, internal lab findings, or functional data^5^.
2. **Genotypic data:** One or more genetic variants and their allele state (heterozygous, homozygous, etc) as found in a patient.
3. **Phenotypic data:** Patient signs and symptoms and structured phenotype descriptors commonly used in variant interpretation and diagnostic assessments, such as, age, sex, race, phenotypic features, etc.
4. **Co-occurrence data**: The presence/absence of other variants that may affect the interpretation of the variant in question (for example, the fact that a patient has a known pathogenic variant in cis with a variant under evaluation).
5. **Segregation data:** Variant tracks with disease in a family, which usually require family history information.

## Appendix 3: Summary table of all coded data (Please see separate attached Excel file.)

## List of Abbreviations

DNA: Deoxyribonucleic acid
EU: European Union
GDPR: General Data Protection Regulation
HCP: Healthcare provider
HIPAA: Health Insurance Portability and Accountability Act
HPO: Human Phenotype Ontology
MME: Matchmaker Exchange
U.S.: United States

